# Effectiveness of drugs for COVID-19 inpatients in Japanese medical claim data as average treatment effects with inverse probability weighted regression adjustment: Retrospective observational study

**DOI:** 10.1101/2023.05.12.23289913

**Authors:** Shingo Mitsushima, Hiromasa Horiguchi, Kiyosu Taniguchi

## Abstract

**Background:** Prior studies have indicated that drugs against coronavirus disease 2019 (COVID-19) such as antiviral drugs, anti-inflammatory drugs, steroid and antibody cocktails are expected to prevent severe COVID-19outcomes and death.

**Object:** We analyzed medical claim data in Japan to assess the effectiveness of drugs againstCOVID-19.

**Method:** We applied an average treatment effect model with inverse probability weighted regression adjustment, to the Medical Information Analysis Databank managed by National Hospital Organization in Japan. The outcome was death during hospitalization. Subjects were all inpatients, inpatients with oxygen therapy, and inpatients with respiratory ventilators, by three age classes: all ages, 65 years old or older, and younger than 65 years old. Data on physical characteristics, underlying diseases, administered drugs, the proportion of mutated strains, and vaccine coverage were used as explanatory variables for logistic regression.

**Result:** Estimated results indicated that only an antibody cocktails (sotrovimab, casirivimab and imdevimab) raised the probability of saving life, even though these drugs were administered in few cases. On the other hand, other drugs might raise the probability of death.

**Discussion:** Results indicated that only antibody cocktails was effective to save life using an average treatment effect model with inverse probability weighted regression adjustment. No other drugs such as remdesivir, dexamethasone, baricitinib and tocilizumab were found to be effective to save life, even in the pseudo-situation of random assignment.

## Introduction

Coronavirus disease 2019 (COVID-19) is an infectious disease that results from severe acute respiratory syndrome coronavirus 2 (SARS-CoV-2). The first case of COVID-19 was detected in December 2019 in Wuhan, China. It subsequently spread worldwide. The drug kinds, which included antiviral drugs, antibody cocktails, and steroid and anti-inflammatory drugs, were developed, leading to decreased incidence of severe COVID-19 and death in Japan [1].

Earlier studies and clinical trials have investigated the effectiveness of drugs used against COVID-19 such as an antiviral drug (remdesivir) [2], antibody cocktails (casirivimab/imdevimab, sotrovimab and tixagevimab/cilgavimab) [3-7], a steroid (dexamethasone) [8], RNA-dependent RNA polymerase inhibitors (molnupiravir and favipiravir) [9-12], protease inhibitors (nirmatrelvir/ritonavir) [9, 13], a JAK inhibitor (ruxolitinib) [14], and anti-inflammatory drugs (baricitinib and tocilizumab) [15-17]. All are expected to prevent severe COVID-19 outcomes and death. For this study, we considered the real-world effectiveness of drugs against COVID-19.

Medical Information Analysis Databank (MIA) is operated by the National Hospital Organization (NHO) in Japan, which provides a database of medical claims from 60 representative NHO hospitals [18]. NHO is one of the biggest hospital organization in Japan, it has about 3.4% of all beds in Japan [19]. We used MIA data to analyze drug effectiveness.

In reality, the decision to administer drugs depends on the patient’s condition: that is not a random assignment. Generally speaking, patients who are more likely to develop severe illness have higher probability of being administered drugs, which implies a negative association between drugs and death because of selection bias in the observational data. Therefore, non-random choice of drug administration is regarded as an estimate of drug effectiveness.

The average treatment effect model with inverse probability weighted regression adjustment might resolve this difficulty statistically but not experimentally [20]. This procedure predicts the likelihood of administration for the drug. Then outcomes were estimated, using logistic regression including dummy variables, whether a drug was administered or not, weighted with the inverse probability of each subject actually belonging to the group estimated by first-step logistic regression for drug administration. It focused more on subjects whose probability of belonging to the group were lower, in other words, less likelihood was regarded as a more nearly random assignment.

Researchers in natural science including medicine can conduct the experiments and thus delete selection bias in the choice of subjects, even though experiments are expensive and require a longer period, usually. Conversely, researchers in social sciences cannot conduct experiments to individual’s choice. Instead of that, they sometimes use the average treatment effect model with propensity score matching, mainly, for the evaluation of programs to control participants choose to join the program.

However, even in medicine, random assignment experiments after launch are difficult. Therefore, situations change after a trial: mutated strains emerge, and vaccine coverage and/or developed treatments might affect drug effectiveness, rendering evaluation impossible. Particularly, mortality is often not used as an outcome for evaluation, even though it is the most important endpoint. Therefore, experiments in medicine for changing situations and mortality might be difficult. To resolve this difficulty, the average treatment effect model was used for evaluation in a changing situation through a statistically pseudo-random assignment experiment. Particularly, it was applied for orthopedic surgery and cardiovascular research [21, 22]. However, these studies used propensity scoring matching method: a simple comparison of outcomes among the treatment group and non-treatment group with almost identical probability to receive treatment. In other words, this method did not control for outcomes other than treatment. It might bias the result by the potential confounder. Therefore, we used another average treatment model with inverse probability weighted regression adjustment. Its second step was weighted regression for the outcome weighted with inverse probability of the first step for whether the patient was treated, or not [23,24]. It can control covariates for outcomes other than whether the patient was treated, or not. Conversely, average treatment effect model with propensity score matching cannot control other condition than whether the patient was treated, or not.

## Materials and Methods

### Data sources

This study used MIA for confirmed inpatients including their age, sex, underlying diseases, hospitalization week, administered drugs against COVID-19, outcome and oxygen therapy and/or use of respiratory ventilator. We had accessed to these data which could identify individual participants.

We used data for vaccine administration published by the Cabinet Secretariat [25]. Moreover, prevalence in mutated strains was referred from a monitoring meeting in Tokyo because MIA included no information about a patient’s vaccine status or sublineage in SARS-CoV-2 [26].

The study period was January 2020 through March 2022, using data recorded as of May 2022, which was the date we conducted this study. The study area was the entirety of Japan.

### Subjects

Subjects were all inpatients who were diagnosed as COVID-19 (U07 in ICD10) from January 2020 through March, 2022. We collected the data on all COVID-19 inpatients in MIA data. Some inpatients who were still hospitalized at the end of study period were excluded. The number of inpatients who were diagnosed as COVID-19 in MIA data moves like 6 waves. These waves were classified from the trough of the prior wave to the trough of the current wave using national data [27]. We defined the 1st wave extended from week 1 of 2020 through week 23 of 2020; the 2nd wave lasted from week 24 of 2020 through week 39 of 2020; the 3rd wave was recorded from week 40 of 2020 through week 8 of 2021; the 4th wave occurred from week 9 of 2021 through week 24 of 2021; the 5th wave was from week 25 of 2021 through week 47 of 2021; and the 6th wave was from week 48 of 2021 through the end of this study.

However, apart from purely medical criteria, the criteria for hospitalization of asymptomatic patients or mild patients who do not require oxygen therapy may be strongly influenced by social conditions, such as lack of medical resources or support for staying and recovering at home. Therefore, we considered not only the inclusion of all subjects but also the limitation of subjects on oxygen therapy or respiratory ventilation.

Note that we did not distinguish some patients with initiation drug before oxygen therapy or respiratory ventilation and patients with initiation drug after oxygen therapy or respiratory ventilation. Because timing of drug initiation, and oxygen therapy or respiratory ventilation using were not available in this study. In other words, patient condition when initiation drug was not accounted for drug effectiveness.

### Definitions of Variables

Physical characteristics: Age and sex.

Underlying diseases: We examined cancer (C00-C90 in ICD10), asthma (J45), chronic obstructive pulmonary disease (COPD) (J44), hypertension (I10), heart failure (I50), and DM (E10).

Pharmaceutical therapy: We examined the effects of an antiviral drug (remdesivir), an antibody cocktails (sotrovimab and casirivimab/imdevimab), a steroid (dexamethasone), and anti-inflammatory drugs (baricitinib and tocilizumab). The antibody cocktails were not divided by drug such as sotrovimab and casirivimab/imdevimab because the number of antibody cocktails was not sufficient. The overall effects of the antibody cocktails were examined. We also collected the number of prescriptions such as RNA-dependent RNA polymerase inhibitors (molnupiravir and favipiravir), protease inhibitors (nirmatrelvir/ritonavir). Remdesivir, antibody cocktails (sotrovimab and casirivimab/imdevimab), dexamethasone, baricitinib, and tocilizumab were the top five drugs which earlier studies had shown as effective.

Vaccine coverage: The complete rate of second dose of vaccine received two weeks prior by age class, as younger than 65 years old (young patients) or 65 years old or older (elderly patients).

Mutated strains: Mutated strains were measured by percentage at one week before admission. Omicron included BA.2 or a later sublineage. Alternatively, we used a dummy variable during the 4th–6th wave instead of the proportion of the mutated strains as an explanatory variable, to check robustness. By this specification, the Alpha variant strain emerged and then dominated in the 4th wave. Similarly, the Delta variant strain emerged and then became dominant in the 5th wave, BA.1 strain emerged and then dominated from the 6th wave.

Outcome: Death during hospitalization.

### Statistical analysis

We conducted estimates separately by drug type: remdesivir, antibody cocktails (sotrovimab, casirivimab and imdevimab), dexamethasone, baricitinib, and tocilizumab.

The estimation procedure was the average treatment effect model with inverse probability weighted regression adjustment. It consists of two steps. The first step to assess whether the patient was administered a type of drug or not was performed through logistic regression on their physical condition, underlying diseases, drugs, vaccine coverage, and prevalence in mutated strains as explanatory variables. The second step was logistic regression for fatality weighted with inverse probability estimated by the first step. Explanatory variables in this step were binomial variables: whether the patients were administered the considered drug or not.

We adopted 5% level as significance level. All statistical analyses were conducted using software (STATA SE 17.0; Stata Corp.).

### Ethical considerations

This study was approved by the Ethics Committee of Mie Hospital (Approval No. 2020-89). Permission to use MIA data was obtained using the NHO (Registration No. 1201003).

## Results

The number of COVID-19 inpatients in this study was 21727. Fig 1 shows the number of COVID-19 inpatients, COVID-19 inpatients with oxygen therapy, COVID-19 inpatients with respiratory ventilation and fatalities due to COVID-19 by the hospitalized week in MIA data. Fig 2 presents the drugs against COVID-19 by the week in which the COVID-19 inpatient administered the drug was admitted to the hospital in MIA data. Fig 1 shows similar trends in COVID-19 inpatients, COVID-19 inpatients with oxygen therapy, COVID-19 inpatients with respiratory ventilation and fatalities due to COVID-19. Fig 2 also shows that all drugs for treating COVID-19 have a similar trend. The most prescribed drugs were dexamethasone, followed by remdesivir, baricitinib, tocilizumab and antibody cocktails (sotrovimab, casirivimab/imdevimab).

**Fig 1.**
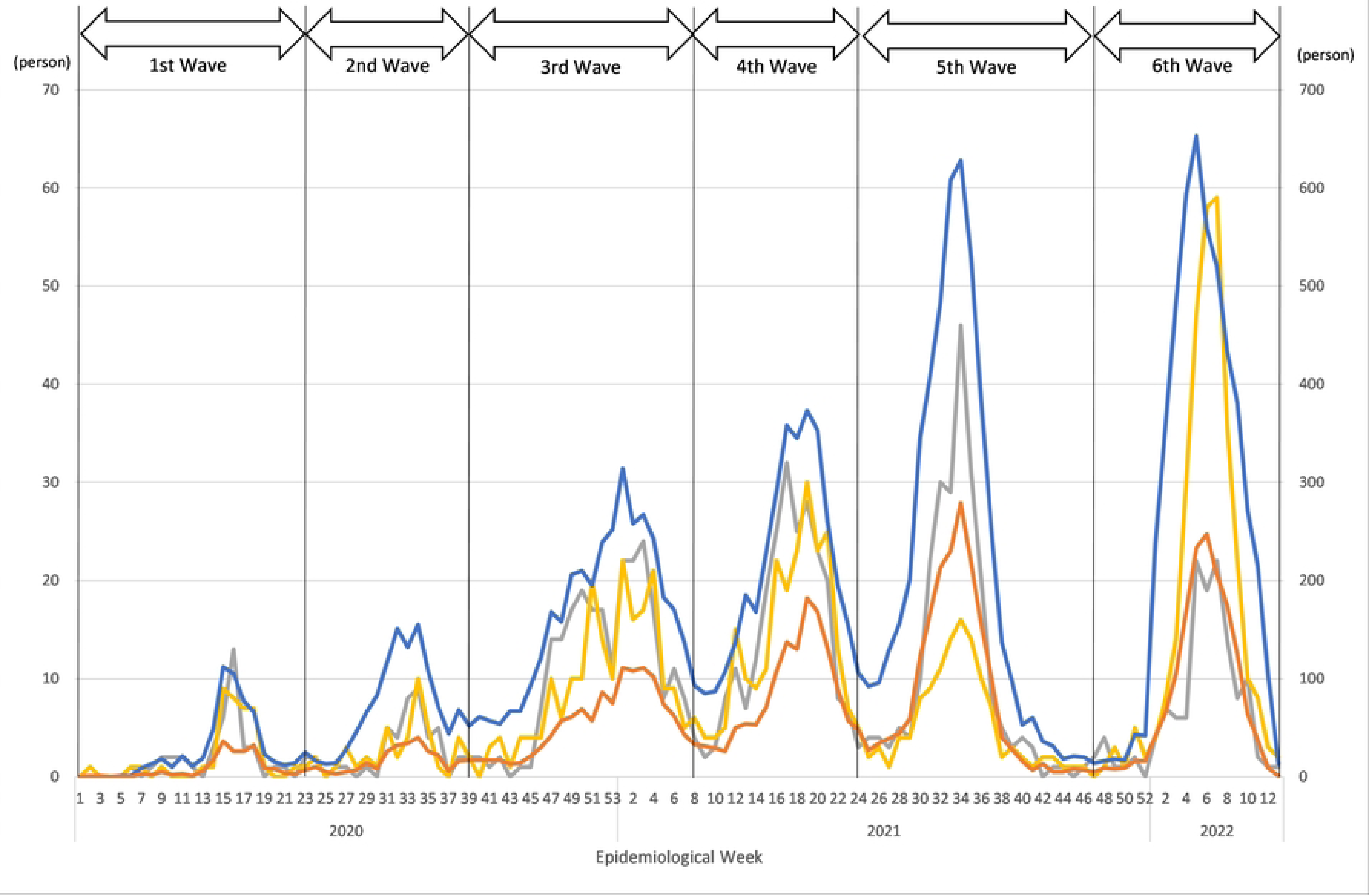
COVID-19 inpatients, COVID-19 inpatients with oxygen therapy, COVID-19 inpatients using respiratory ventilation and fatalities attributed to COVID-19 by the hospitalized week in MIA data. Notes: Blue line and orange line show the numbers of COVID-19 inpatients and inpatients with oxygen therapy respectively in MIA data (right scale). Gray line and yellow line show the numbers of COVID-19 inpatients with respiratory ventilator and fatalities due to COVID-19 respectively in MIA data (left scale).

**Fig 2.**
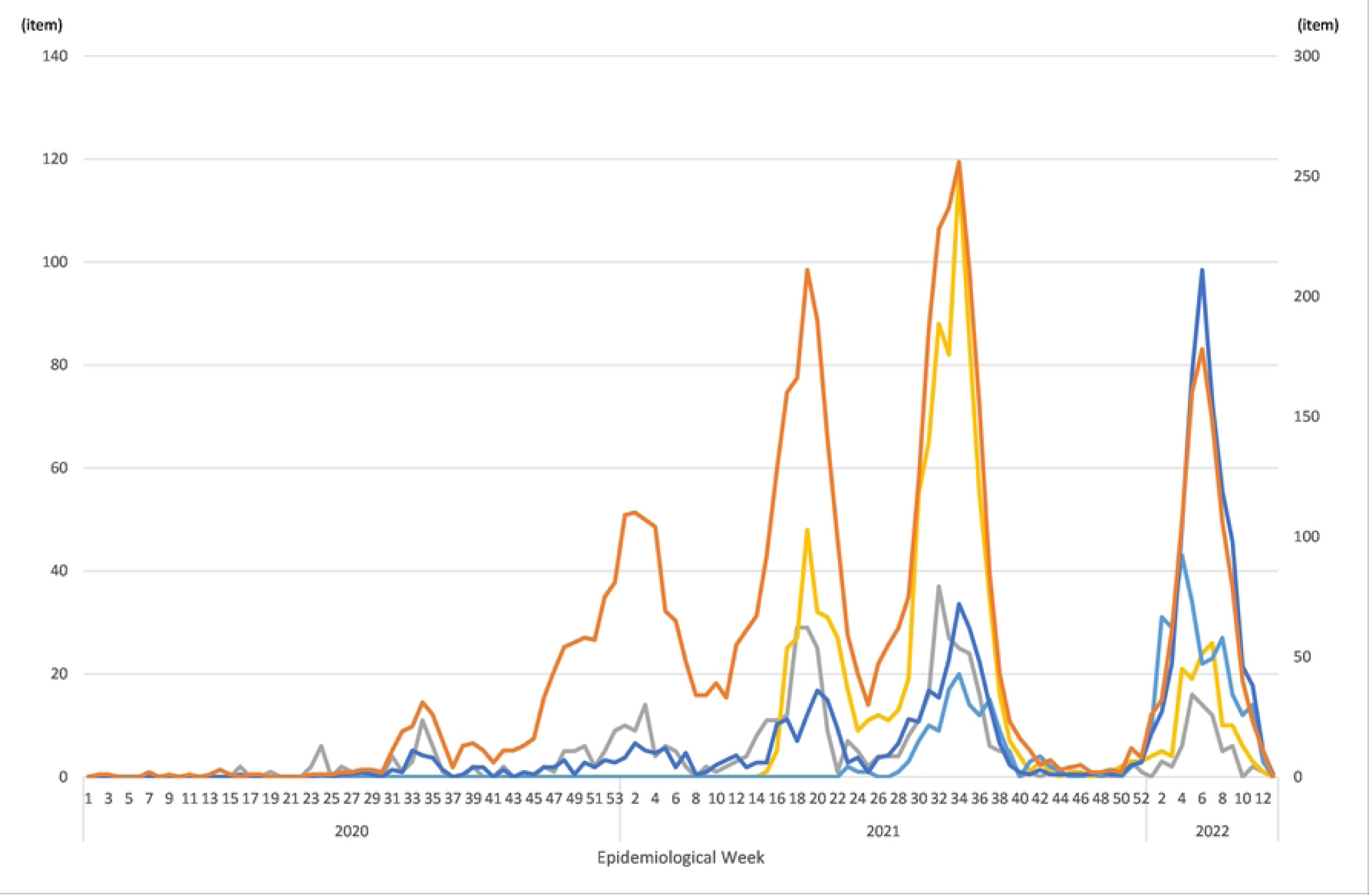
Drugs for treating COVID-19 in MIA data. Notes: Blue line and orange line show the numbers of remdesivir and dexamethasone respectively in MIA data (left scale). Gray line, yellow line and light blue line show the numbers of tocilizumab, baricitinib and antibody cocktails (sotrovimab, casirivimab/imdevimab) respectively in MIA data (right scale).

Table 1 shows the characteristics 21727 inpatients. The mean of their age was 54.3. The proportion of sex of male and female was almost same. The most common underlying disease was diabetes mellitus followed by hypertension, cancer, heart failure, asthma, and COPD. Dexamethasone was the most prescribed drug, followed by remdesivir, baricitinib, tocilizumab, and antibody cocktails. Fig 3 presents proportions of mutated strains (Alpha, Delta and Omicron variants) in Tokyo and vaccination coverage with two and three doses in Japan. The predominant variant changed from Alpha to Delta in July 2021 and from Delta to Omicron in December 2021. Vaccination coverage with two doses has increased since May 2021 and reached 80% by the end of 2021.

**Fig3.**
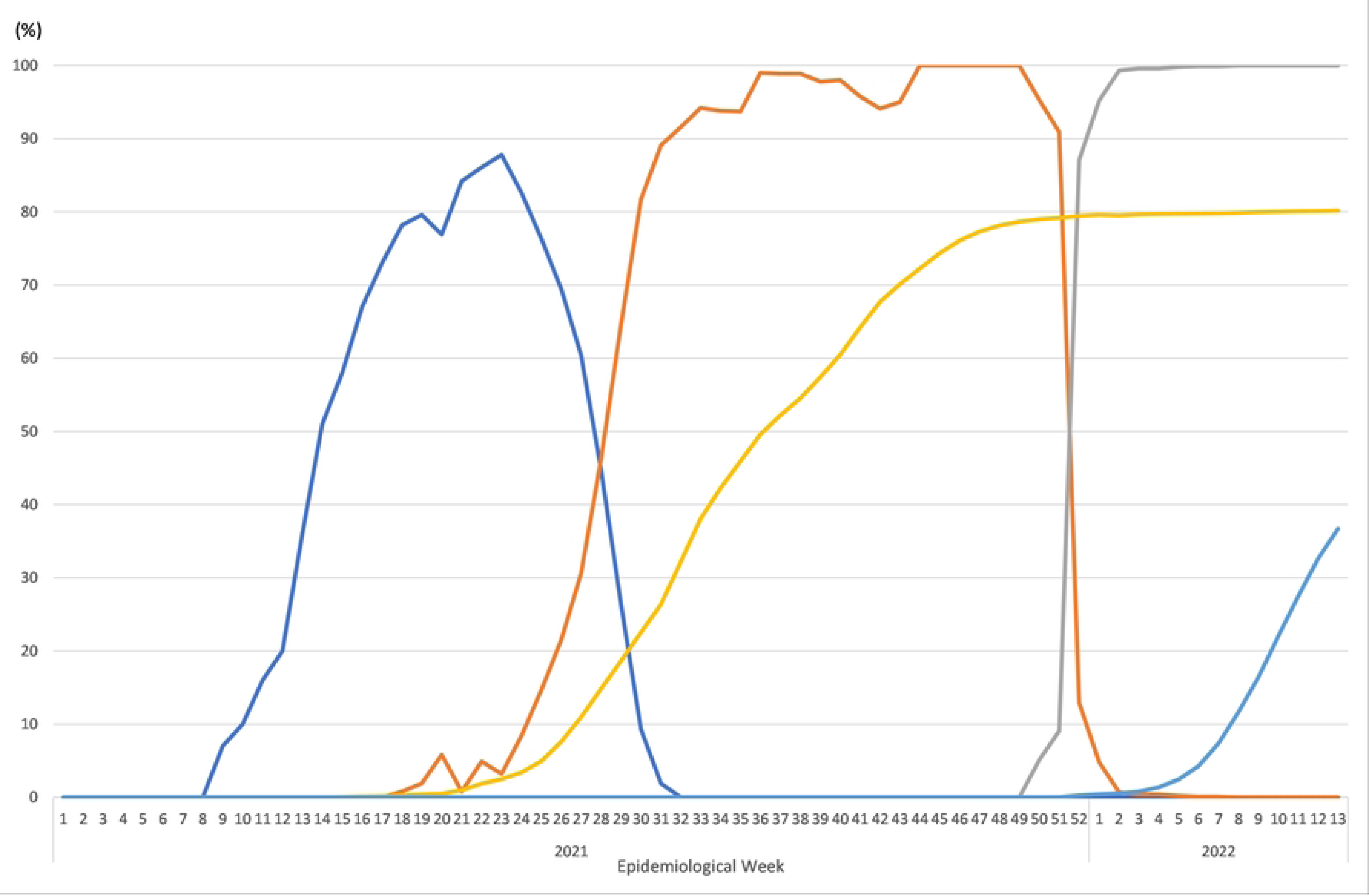
Proportions of Alpha, Delta and Omicron variants in Tokyo, in addition to vaccination coverage with two and three doses in Japan. Notes: Blue, orange, and gray lines respectively represent the proportions of the Alpha, Delta and Omicron variants. Data of the proportions of the Alpha and Delta variants were published by the Cabinet Secretariat. Yellow and light blue lines respectively represent vaccination coverage with two and three doses in the entire Japanese population. Data of the vaccination coverage were published by the monitoring meeting in Tokyo.

**Table 1.** Characteristics of subjects.

| Characteristics | All inpatients |  |
| --- | --- | --- |
|  | n | % |
| Age |  |  |
| Mean (SD) | 54.3 (25.6) |  |
| Sex |  |  |
| Female | 10083 | 46.4 |
| Underlying diseases |  |  |
| Cancer | 1036 | 4.8 |
| Hypertension | 2820 | 13 |
| Diabetes mellitus | 3241 | 14.9 |
| Heart failure | 825 | 3.8 |
| Asthma | 575 | 2.6 |
| COPD | 217 | 1.0 |
| Drugs against COVID-19 |  |  |
| Remdesivir | 1910 | 8.8 |
| Dexamethasone | 6041 | 27.8 |
| Tocilizumab | 562 | 2.6 |
| Baricitinib | 1087 | 5.0 |
| Antibody cocktails | 525 | 2.4 |

Note: Range of age was 0-108.
Abbreviation: SD: standard deviation, COPD: chronic obstructive pulmonary disease

Table 2 presents the estimation results. Overall, four drugs except for antibody cocktails (sotrovimab, casirivimab/imdevimab), remdesivir, dexamethasone, baricitinib, and tocilizumab had positive treatment effects among those found to be significant. That finding might indicate that these drugs reduce the probability of saving life. Conversely, antibody cocktails had negative treatment effects which indicated a contribution to saving life. Only four pairs were identified to estimate parameters; other combinations were not.

**Table 2.** Estimation Results of Average Treatment using Inverse Probability Weighted Regression Adjustment.

| Age class | All | 65 years old or older | Younger than 65 years old |
| --- | --- | --- | --- |

|  | difference | *p* value | difference | *p* value | difference | *p* value |
| --- | --- | --- | --- | --- | --- | --- |
| Remdesivir |  |  |  |  |  |  |
| Proportion |  |  |  |  |  |  |
| All | 0.042 | 0.000 | 0.099 | 0.000 | 0.007 | 0.331 |
| Oxygen therapy | 0.043 | 0.000 | 0.070 | 0.000 | 0.002 | 0.746 |
| Respiratory ventilator | 0.034 | 0.370 | 0.071 | 0.160 | 0.001 | 0.987 |
| Period |  |  |  |  |  |  |
| All | 0.040 | 0.000 | 0.094 | 0.000 | 0.006 | 0.335 |
| Oxygen therapy | 0.041 | 0.000 | 0.067 | 0.000 | 0.002 | 0.765 |
| Respiratory ventilator | 0.031 | 0.419 | 0.063 | 0.194 | -0.029 | 0.415 |
| Dexamethasone |  |  |  |  |  |  |
| Proportion |  |  |  |  |  |  |
| All | 0.039 | 0.000 | 0.090 | 0.000 | 0.004 | 0.017 |
| Oxygen therapy | 0.032 | 0.000 | 0.056 | 0.000 | 0.002 | 0.684 |
| Respiratory ventilator | -0.006 | 0.833 | -0.017 | 0.670 | 0.005 | 0.889 |
| Period |  |  |  |  |  |  |
| All | 0.039 | 0.000 | 0.088 | 0.000 | 0.004 | 0.022 |
| Oxygen therapy | 0.031 | 0.000 | 0.053 | 0.000 | 0.002 | 0.661 |
| Respiratory ventilator | -0.005 | 0.858 | -0.013 | 0.735 | 0.005 | 0.891 |
| Tocilizumab |  |  |  |  |  |  |
| Proportion |  |  |  |  |  |  |
| All | 0.114 | 0.000 | 0.255 | 0.000 | 0.025 | 0.010 |
| Oxygen therapy | 0.127 | 0.000 | 0.219 | 0.000 | N/A | N/A |
| Respiratory ventilator | 0.098 | 0.027 | N/A | N/A | -0.011 | 0.776 |
| Period |  |  |  |  |  |  |
| All | 0.117 | 0.000 | 0.259 | 0.000 | 0.025 | 0.010 |
| Oxygen therapy | 0.129 | 0.000 | 0.219 | 0.000 | N/A | N/A |
| Respiratory ventilator | 0.105 | 0.016 | N/A | N/A | -0.030 | 0.441 |
| Baricitinib |  |  |  |  |  |  |
| Proportion |  |  |  |  |  |  |
| All | 0.072 | 0.002 | 0.371 | 0.000 | -0.001 | 0.766 |
| Oxygen therapy | 0.107 | 0.006 | 0.316 | 0.000 | -0.004 | 0.519 |
| Respiratory ventilator | 0.281 | 0.000 | 0.376 | 0.000 | N/A | N/A |
| Period |  |  |  |  |  |  |
| All | N/A | N/A | N/A | N/A | N/A | N/A |
| Oxygen therapy | N/A | N/A | N/A | N/A | N/A | N/A |
| Respiratory ventilator | N/A | N/A | N/A | N/A | N/A | N/A |
| Sotrovimab or<br>casirivimab/imdevimab |  |  |  |  |  |  |
| Proportion |  |  |  |  |  |  |
| All | -0.042 | 0.000 | -0.097 | 0.000 | -0.001 | 0.878 |
| Oxygen therapy | -0.074 | 0.000 | -0.125 | 0.000 | N/A | N/A |
| Respiratory ventilator | N/A | N/A | N/A | N/A | N/A | N/A |
| Period |  |  |  |  |  |  |
| All | N/A | N/A | N/A | N/A | N/A | N/A |
| Oxygen therapy | N/A | N/A | N/A | N/A | N/A | N/A |
| Respiratory ventilator | N/A | N/A | N/A | N/A | N/A | N/A |

**Notes:** Yellow denotes significance. “N/A” denotes not available.

Overall, all age and older patients had more significant estimators than younger patients had. Moreover, few significant estimates were found when the subjects were limited to patients who had used a respiratory ventilator. Only for tocilizumab in all ages were these found to be significantly positive. Only four parameters were identified for estimation for antibody cocktails, even though these were all negative and significant.

Estimation results for mutated strains were defined as the proportion, which was similar with that defined as the period, except for antibody cocktails. Among the results obtained for antibody cocktails, there is no pair to estimate parameters.

## Discussion

Results showed that antibody cocktails might contribute to saving life consistently. Our earlier studies, simple logistic regression and estimation using propensity score matching, yielded similar results [28,29]. However, estimation results of propensity score matching method for tocilizumab and baricitinib were, respectively, positive and negative. In this study, we obtained consistent estimated results for tocilizumab and baricitinib. These results indicated that tocilizumab and baricitinib might not contribute to saving life. Even for antibody cocktails, the effects for younger or more severe patients who used respiratory ventilators were not confirmed. Prior study showed that casirivimab/imdevimab, one of antibody cocktails, reduced viral load [3]. Antibody cocktails were usually administered for mild COVID-19 patients, that is, they were administered before viral load increased significantly. Therefore, they might be more effective to COVID-19 compared to drugs used for severe COVID-19 patients. The small sample for the younger patients or severe patients might be also another reason.

These counterintuitive findings, except for those for antibody cocktails, which are inconsistent with results obtained from earlier studies of the effectiveness of remdesivir and dexamethasone [2, 8], might result from worse matching at the first step. In addition, endogeneity or selection bias might not be controlled well in the decisions to use drugs. Drugs to be administered against COVID-19 depend on the severity of illness in patients. For instance, an earlier report described dexamethasone as effective for severe COVID-19, but as ineffective against mild or moderate COVID-19 [8]. Actually, more than antibody cocktails, four other drugs were reportedly more effective for severely ill patients [2-4, 8]. To resolve this apparent contradiction, more information indicating severity might be necessary to make more precise the first step estimation. Moreover, we did not use test results such as those for blood pressure, BMI, or oxygen saturation. Such information must be included in the first step. Inclusion of such data remains as a challenge for future research. The results of antibody cocktails included many “not available”, although drug effectiveness was confirmed in matched cases. This result might be attributed to the small number of antibody cocktails used in patients. In Japan, remdesivir, dexamethasone, baricitinib, casirivimab/imdevimab, sotrovimab, and tocilizumab were approved in May 2020, July 2020, April 2021, July 2021, September 2021, and January 2022 respectively [30]. More recently approved drugs are likely to be smaller samples. Accumulating data may resolve this shortcoming. We also have to pay attention to apply the results of this study to all COVID-19 patients because the targets of this study were all inpatients.

## Limitations

First, we estimated drug effectiveness separately. However, the choice of drug was not actually independent. Therefore, intercorrelation among drugs should be incorporated into the estimation model.

Second, because MIA is a database of medical claims, data from the most recent weeks may change during next few months. For this study, data were collected and analyzed from January, 2020 through the end of March, 2022, as of May, 2022. If the study period is extended, the data and its estimation may differ over time.

Third, the drugs examined in this study were not necessarily administered to the severest patients. For example, antibody cocktails were often administered to mild patients. However, to simplify the analysis, we did not consider the combination of drugs or change from one considered drug to another drug. This choice might introduce some bias in the results. This process or pattern of drug administration may be important in evaluating drug effectiveness.

Fourth, due to data availability, it was not possible to take into account the timing of drug initiation, and oxygen therapy or respiratory ventilation to estimate drug effectiveness. If this information were available, it would be possible to distinguish between patients who were administered drugs before oxygen therapy or respiratory ventilation and patients who were administered drugs after oxygen therapy or respiratory ventilation. It might be useful for more careful estimation to drug effectiveness.

## Conclusion

The obtained results demonstrated that antibody cocktails for all or older patients might contribute to the saving of life. However, more information, including test results, is necessary for better matching and achievement of definitive conclusions.

## Data availability

The data in this study are available from the National Hospital Organization, but the availability of these data is restricted because of privacy reasons. These data were used under license for this study and are therefore not available to the public. Data are available from the authors with permission of the Ethics Committee and National Hospital Organization.

## Acknowledgements

We acknowledge Mr. Masaya Nakadera and Mr. Masato Koizumi, who prepared the database, and thank all hospitals for submitting patient data.

